# Combining Sodium MRI, Proton MR Spectroscopic Imaging and Intracerebral EEG in Epilepsy

**DOI:** 10.1101/2022.08.03.22278332

**Authors:** Mikhael Azilinon, Julia Scholly, Wafaa Zaaraoui, Samuel Medina Villalon, Patrick Viout, Tangi Roussel, Mohamed Mounir El Mendili, Ben Ridley, Jean-Philippe Ranjeva, Fabrice Bartolomei, Viktor Jirsa, Maxime Guye

## Abstract

Whole brain ionic and metabolic imaging has potential as a powerful tool for the characterization of brain diseases. In this study we combined sodium MRI (^23^Na MRI) and ^1^H-MR Spectroscopic Imaging (^1^H-MRSI) and compared ionic/metabolic changes probed by this multimodal approach to intracerebral stereotactic-EEG (SEEG) recordings.

We applied a multi-echo density adapted 3D projection reconstruction pulse sequence at 7T (^23^Na MRI) and a 3D echo planar spectroscopic imaging sequence at 3T (^1^H-MRSI) in 19 patients suffering from drug-resistant focal epilepsy who underwent presurgical SEEG. We investigated ^23^Na MRI parameters including total sodium concentration (TSC) and the sodium signal fraction associated of with the short component of T_2_* decay (*f*), alongside the level of metabolites N-acetyl aspartate (NAA), choline compounds (Cho) and total creatine (tCr). All measures were extracted from spherical regions of interest (ROIs) centered between two adjacent SEEG electrode contacts and z-scored against the same ROI in controls.

Group comparison showed a significant increase in *f* only in the epileptogenic zone (EZ) compared to controls and compared to patients propagation zone (PZ) and non-involved zone (NIZ). TSC was significantly increased in all patients’ regions compared to controls. Conversely, NAA levels were significantly lower in patients compared to controls, and lower in the EZ compared to PZ and NIZ. Multiple regression analyzing the relationship between sodium and metabolites levels revealed significant relations in PZ and in NIZ but not in EZ.

Our results are in agreement with the energetic failure hypothesis in epileptic regions associated with widespread tissue reorganization.

## 1. Introduction

Recent advances in neuroimaging have challenged the concept of focal epilepsy as a brain disorder strictly limited to the regions responsible for seizure generation and propagation (Larivière et al., 2021). Various MRI modalities (i.e. structural, functional, metabolic) have consistently demonstrated structural and functional alterations that extend beyond the epileptogenic regions and affect areas non-involved in ictal discharges or interictal epileptiform activity. These complex changes affect both structure and function at different spatial and temporal scales and can reflect seizure-induced alterations, neuronal plasticity as well as the underlying etiology. These factors, and the precise anatomical location of generators of epileptiform activity, are subject to variation even between individuals with the same epileptic syndrome. Thus, a systematic comparison of imaging data with the gold standard of electrophysiological data derived from intracerebral stereoelectroencephalography (SEEG) recording is essential to test the potential contribution of any imaging modality towards better definition of the epileptogenic zone (EZ) in patients suffering from drug-resistant focal epilepsy who are candidates for surgery. Moreover, the need to characterize the variable manifestations of pathology suggest a clear need to exploit multimodal imaging, combining metrics from different modalities. Insight into alterations of ionic homeostasis and metabolic function can be probed by sodium (^23^Na) MRI and ^1^H-MR spectroscopic imaging (MRSI), respectively.

^23^Na-MRI provides a unique opportunity to non-invasively image sodium signal in the brain (Madelin et al., 2014). To date, the only ^23^Na-MRI study performed in a group of human focal epilepsy, has demonstrated an increase of the total sodium concentration (TSC) in patients’ brain compared to controls, which was greater in the epileptogenic zone (EZ) compared to the propagation zone (PZ) and the non-involved regions (NIZ) (Ridley et al., 2017). The usefulness of TSC as a potential epileptogenicity marker may remain limited due to its limited specificity, as it likely reflects different underlying phenomena at the cellular level, such as changes in intracellular sodium concentration, changes in extracellular volume and cells density and/or organization among others.

For ^23^Na-MRI the increase in signal to noise available at 7T allows novel approach based on a 3D-multi-echoes density-adapted radial sequence which exploits the biexponential T_2_* decay of the ^23^Na MR signal (Ridley et al., 2018). This approach permits a multiparametric investigation of variation in T_2_* decay behavior related to the quadrupolar interactions of the 3/2 spin of ^23^Na with the electric field gradient of surrounding molecules (Rooney & Springer, 1991), as an indicator of tissue organization and molecular environment. The biexponential fit model can be used as a probe to determine the motional regimes of sodium nuclei within the surrounding environment. Thus, by quantifying the sodium signal fraction with the short T_2_* decay component (*f*) this approach may offer a more relevant metric for studying tissue alterations and potentially provide a better link between sodium homeostasis and neuronal excitability in human epilepsy. In the present study, we implemented this multiparametric approach of sodium MRI for the first time in the assessment of profiles within and outside the epileptogenic and propagation networks in focal drug-resistant epilepsy.

As a further objective, we assessed metabolic alterations accompanying sodium concentration changes. To do so, we explored metabolic status by using whole-brain ^1^H-Echo Planar spectroscopic imaging (^1^H-EPSI) in the same subjects at 3T. Three main metabolites were quantified : (i) N-acetyl Aspartate (NAA) reflecting neuronal viability, mitochondrial dysfunction or neuronal loss (Moffett et al., 2013; Stefano et al., 1995), (ii) Choline compounds (Cho) reflecting membrane turnover and inflammatory processes (Achten, 1998; Urenjak et al., 1993), and (iii) total-creatine compounds (tCr) reflecting intracellular energy states, and energy-dependent systems in the brain (Kreis et al., 1992; Kreis & Ross, 1992), and considered as a cellularity index (Kreis et al., 1993). Importantly, NAA has been consistently shown to be decreased in the epileptic brain, particularly in the EZ and PZ, compared to non-involved regions, and is thus considered a potential epileptogenicity marker in focal epilepsy (Guye et al., 2002, 2005; Hugg et al., 1993; Kuzniecky et al., 1998; Lundbom et al., 2001; Simister et al., 2002). Furthermore, it has been hypothesized that the observed increase in total sodium concentration would mainly reflect energetic failures due to mitochondrial dysfunction affecting the Na^+^/K^+^ pump activity (Ridley et al., 2017; Stys et al., 1992). Thus, measuring both sodium and NAA in the same regions provides clues with regard to the mitochondrial defect hypothesis (Donadieu et al., 2019; Paling et al., 2011). However, the use of this metabolite has been limited by poor resolution and spatial coverage of routinely performed ^1^H-MRSI, as well as by its insufficient specificity. In this study, we benefited from the whole brain coverage with a relatively high spatial resolution allowing comparison between multimodal MRI and electrophysiological metrics.

Therefore, through this trimodal approach, we aimed to characterize ionic and metabolic changes within epileptogenic networks in comparison with electrophysiologically normal appearing brain networks. For this purpose, we analyzed differences in ^23^Na-MRI (TSC and *f*) and ^1^H-MRSI (Cho, NAA and tCr) metrics between patients and controls as well as between regions of interest (ROI) defined by quantitative SEEG signal analysis (Figure 1). We then investigated the association between ^23^Na-MRI and ^1^H-MRSI metrics in EZ, PZ and NIZ in order to link the homeostatic and metabolic mechanisms to the SEEG recorded electrical alterations.

**Figure 1:**
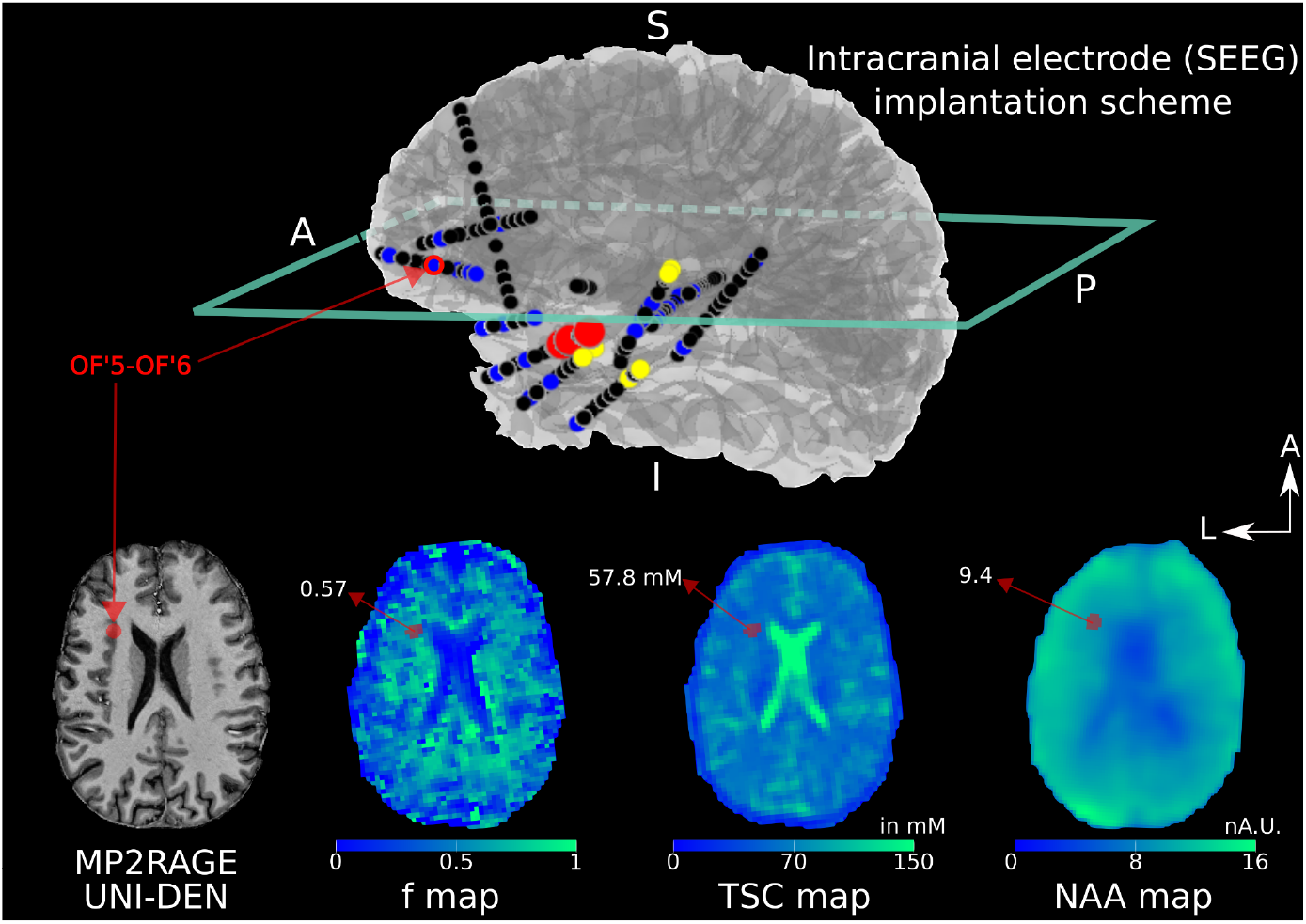
3D representation of an example of SEEG electrode implantation scheme (top) and f, TSC and NAA maps slices corresponding to the transversal green section on the 3D view. Red spherical ROIs correspond to regions belonging to the epileptogenic zone (EZ), yellow ROIs correspond to regions belonging to the propagation zones (PZ), blue ROIs correspond to regions non-involved by electrical abnormalities (NIZ) and black ROIs correspond to regions not explored in this study. Black spheres correspond to regions excluded from the analyses based on the SEEG signal. The red dot in the transverse planes (i.e. T1w image and f, TSC and NAA maps) represents the ROI corresponding to electrode OF’ (left orbitofrontal location) contact 5 and 6 (or bipolar contact 5-6). OF’5-OF’6 ROI is circled in red in the implantation scheme. Red arrows are pointing to the mean ROI value for each map.

## 2. Materials and Methods

### 2.1. Subjects

Among all patients who underwent stereotactic intracerebral EEG (SEEG) recording in the context of presurgical evaluation for drug-resistant focal epilepsy at our center between January 2017 and February 2020, 19 consecutive patients with available 3D ^1^H-MRSI and ^23^Na-MRI were retrospectively included (Table 1). All patients had detailed non-invasive presurgical evaluation including medical history, neurological examination, neuropsychological assessment, ^18^F-FDG-PET, high-resolution structural 7T and 3T MRI, and a long-term scalp-video-EEG. All these steps were necessary for patient enrollment. The SEEG was indicated in all patients to localize the EZ and to precisely determine its relation with eloquent areas. SEEG was performed as a part of the routine clinical management in line with the French national guidelines on stereoelectroencephalography (SEEG) (Isnard et al., 2018). SEEG implantation was planned individually for each patient, according to anatomo-electro-clinical hypotheses about the localization of the EZ based on non-invasive investigations. All SEEG explorations were bilateral and systematically sampled temporal, insular, frontal, and parietal regions of at least one hemisphere. Follow-up information was collected from a review of the medical records.

**Table 1.** Patient clinical demography.

| Patient | Gender | Epilepsy duration (y) | Seizure frequency (per month) | Epilepsy type | Side | 3T/7T anatomical MRI | Surgical procedure | Engel class | Histology | <sup>23</sup>Na-MRI/<sup>1</sup>H-MRSI |
| --- | --- | --- | --- | --- | --- | --- | --- | --- | --- | --- |
| 1 | F | 14 | 2 | Temporal latero-mesial | L | Normal | TC, SR | II | FCD1 | Both |
| 2 | F | 26 | 20 | Temporo-parietal mesial | R | R posterior cingular FCD | TC, VNS, contra-indication for SR | NA | NA | Both |
| 3 | M | 31 | 2 | Insulo-parietal | L | Normal | TC, VNS contra-indication for SR | NA | NA | Both |
| 4 | F | 9 | 4 | Temporal mesial | R | Normal | TC, SR (ATL) | I | FCD3a | Both |
| 5 | F | 27 | 3 | Temporo-insular | L | Normal | TC, SR | I | FCD1 | Both |
| 6 | M | 26 | 120 | Temporal mesial | L | Normal | TC, SR | II | slight gliosis | Both |
| 7 | M | 11 | 8 | Temporal mesial | L | Normal | TC, SR | II | FCD1b | Both |
| 8 | M | 12 | 4 | Temporo-insular | L | Normal | TC, contra-indication for SR | NA | NA | Both |
| 9 | M | 24 | 10 | Temporal lateral | L | Normal | TC, SR | III | slight gliosis | Both |
| 10 | F | 9 | 30 | Temporal mesial | R | Normal | TC, SR (ATL) | I | FCD1 | Both |
| 11 | F | 29 | 60 | Orbitofronto-insular | L | L orbitofrontal FCD | TC, SR | III | FCD2a | Both |
| 12 | F | 10 | 30 | L parietal mesial<br>Bilateral temporal mesial | R&L | L posterior cingular NDT | TC, LITT (aiming NDT) | III | NA | Both |
| 13 | F | 18 | 100 | Bilateral temporal mesial,<br>R insulo-operculo-frontal | R>L | Normal | TC, contra-indication for SR | NA | NA | Both |
| 14 | M | 4 | 5 | Temporal latero-mesial | L | Normal | TC, SR | I | FCD2a | <sup>23</sup>Na-MRI |
| 15 | F | 47 | 12 | Temporo-parietal | L | L perisylvian PMG, schizencephaly, temporo-parietal PNH, R posterior PNH | TC, contra-indication for SR | NA | NA | <sup>1</sup>H-MRSI |
| 16 | M | 26 | 12 | Temporal mesial | L | Normal | TC, patient refused SR | NA | NA | <sup>1</sup>H-MRSI |
| 17 | F | 10 | 5 | Bilateral temporal mesial | R&L | Normal | TC, DBS, contra-indication for SR | NA | NA | Both |
| 18 | M | 10 | 12 | Insulo-opercular | L | Normal | TC, seizure free | NA | NA | Both |
| 19 | M | 7 | 60 | Temporal mesial | R | Normal | TC, SR (ATL) | III | FCD1c | Both |

*Abbreviations:* ATL, anterior temporal lobectomy; DBS, deep brain stimulation; DNET, dysembryoplastic neuroepithelial tumor; FCD, focal cortical dysplasia; L, left; LITT, laser-guided interstitial thermal therapy; NA, not applicable; NDT, neuro-developmental tumor; PMG, polymicrogyria; PNH, periventricular nodular heterotopia; R, right; SR, surgical resection; TC, thermocoagulation; VNS, vagus nerve stimulation.

MRI data were always acquired before SEEG implantation. After quality checks (see below), seventeen spectroscopic ^1^H-MRSI datasets, and fifteen ^23^Na-MRI datasets were used for further analysis. Thirteen out of the nineteen patients fulfilled data quality in sufficient ROIs for both modalities. For ^1^H-MRSI, we used a control database of 25 healthy controls (HC; mean age 30.5 ± 9.7 years, range 20-60 years, 14 women). For ^23^Na-MRI, we used a control database of 18 HC for (mean age 30.5 ± 8.36 years, range 21-54 years, 10 women). Participants provided informed consent in compliance with the ethical requirements of the Declaration of Helsinki and the protocol was approved by the local Ethics Committee (Comité de Protection des Personnes sud Méditerranée 1).

### 2.2. MRI Acquisitions

The protocol was conducted for all subjects on the two same MR scanners. The Spectroscopic imaging protocol on a 3-Tesla Magnetom Verio MR system (Siemens, Erlangen, Germany) and the ^23^Na MRI protocol on a whole-body 7-Tesla Magnetom Step 2 MR system (Siemens, Erlangen, Germany).

At 3T, ^1^H-MRI and ^1^H-EPSI were performed with a thirty-two channel phased-array head coil and included a sagittal high resolution 3D-MPRAGE protocol (TE/TR/TI = 3/2300/900 ms, 160 sections, 256×256 mm², FOV 256×256 matrix, resolution = 1 mm³). Whole brain 3D ^1^H-EPSI was acquired as described in (Lecocq et al., 2015) using two axial acquisitions with two different orientations that are the AC-PC plane and the AC-PC + 15° plane (TE/TR/TI = 20/1710/198 ms, nominal voxel size = 5.6 × 5.6 × 10 mm^3^, FOV = 280 × 280 × 180 mm^3^, flip angle = 73°, 50 × 50 × 18 k-space points, GeneRalized Autocalibrating Partial Parallel Acquisition (GRAPPA) factor = 2, acquisition time ≈ 17 minutes). The two angles of EPSI orientations were chosen in order to obtain good quality spectra on as large brain area as possible on at least one acquisition with a reasonable angulation to permit accurate automatic normalization procedure.

At 7T, a high-resolution proton MRI 3D-MP2RAGE (TR = 5000 ms/TE = 3 ms/TI1 = 900 ms/TI2 = 2750 ms, 256 slices, 0.6 mm isotropic resolution, acquisition time = 10 min) was obtained using a 32-element (32Rx/1Tx) ^1^H head coil (Nova Medical). ^23^Na-MRI was acquired using a dual-tuned ^23^Na/ ^1^H QED birdcage coil and a multi-echo density adapted 3D projection reconstruction pulse sequence (TR = 120 ms, 5000 spokes, 384 radial samples per spoke, 3 mm nominal isotropic resolution, 24 echoes (8 per run with 3 runs, acquisition time 10 min per run (30 min in total)). Three dimension ^23^Na MRI volumes were obtained at 24 different TEs ranging from 0.2 ms to 70.78 ms (Run 1: 0.2 - 9.7 - 19.2 - 28.7 - 38.2 - 47.7 - 57.2 - 66.7 ms; Run 2: 1.56 - 11.06 - 20.56 - 30.06 - 39.56 - 49.06 - 58.56 - 68.06 ms; Run 3: 4.28 - 13.78 - 23.28 - 32.78 - 42.28 - 51.78 - 61.28 - 70.78 ms). This ensured a sufficient number and distribution of TEs while taking into account the 5 ms readout of the sequence, especially for measuring ^23^Na signal with short T_2_*. For quantitative calibration of brain sodium concentrations we used as external reference six tubes (80mm length, 15mm diameter) filled with a mixture of 2% agar gel and sodium at different concentrations: two tubes at 25mM, one at 50mM, two at 75mM and one at 100mM. Tubes were positioned in the field of view in front of the subject’s head and maintained using a cap.

### 2.3. MRI Data Processing

Three dimension ^1^H-EPSI images were post-processed with the Metabolite Imaging and Data Analyses System (MIDAS, Trac, MRIR, Miami) (Maudsley et al., 2006). This software ensures B_0_ map correction, lipid suppression, tissue volume fraction through T_1_ segmentation, spectral fitting, exclusion of outlier voxels based on Cramer Rao lower bounds (CRLB) and signal normalization with the interleaved water signal acquired. MIDAS provided AC-PC and AC-PC 15° oriented maps, including metabolite maps (NAA, Cho, tCr), quality maps, CRLB maps and linewidth maps among others that were used in further processing and quality check steps. In this study we only analyzed NAA, tCr and Cho maps because m-Ino (Myo-Inositol) and Glx (for glutamate, glutamine and glutathione) maps did not fulfill CRLB criteria in the majority of the ROIs (Figure 2). ROI selection process is detailed in the next paragraph. The maps that we used for the ROIs signal extraction are the average maps of realigned AC-PC and AC-PC 15° oriented maps (Donadieu et al., 2019; Lecocq et al., 2015) (SPM12, (*Statistical Parametric Mapping: The Analysis of Functional Brain Images - 1st* Edition, n.d.)).

**Figure 2:**
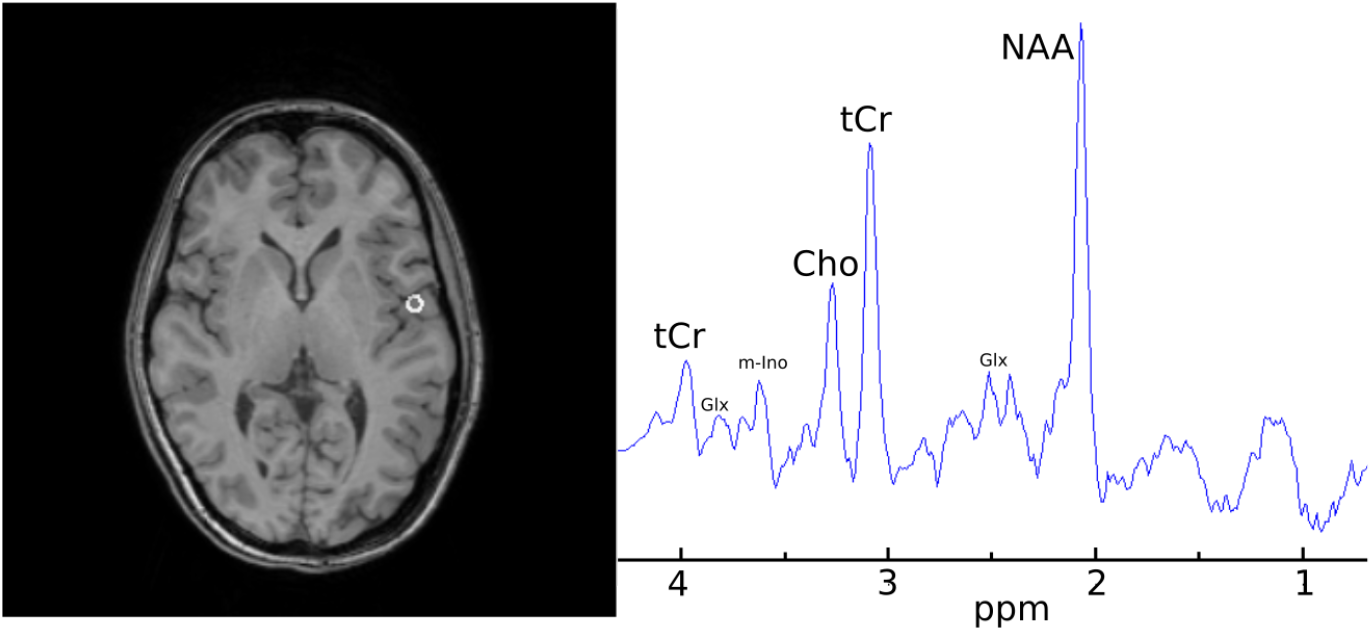
Spectrogram from a healthy control cortical voxel. Here we highlighted the three metabolites we focused on in this study, namely total Choline (Cho), N-Acetyl Aspartate (NAA) and total creatine (tCr). Glx: glutamate, glutamine and glutathione; m-Ino: myo-Inositol.

Metabolic profiles of each ROI (for details on ROI definition see section 2.6) were determined by extracting water signal normalized values (arbitrary unit) from corrected quantitative NAA, tCr and Cho maps derived from MIDAS. We applied quality assurance criteria for spectra in each ROI based on a combination of Cramer-Rao minimum variance bound (CRLB < 15%, or CRLB lower than half of all HC), and water peak linewidth (<16 Hz) thresholds. Supplementary Fig.1. illustrates the spectra that are in the permitted range or not. If, after ROIs rejection in a patient, there was no ROI in EZ, the patient was entirely discarded from further analyzes.

The ^23^Na-MRI data were processed according to (Grimaldi et al., 2021) using a homemade adjusted procedure. After brain extraction on ²³Na-MRI images (ANTS, (Avants et al., 2011)), a denoising filter was applied to the resulting ^23^Na-MRI volumes (Aja-Fernandez et al., 2008; Rajan et al., 2010). The first echo time (TE) volumes from ^23^Na MRI were used as reference for coregistration of the other TE volumes to correct for potential motion between acquisitions. Hand drawn ROIs were placed in the center of each agar tube to extract signal intensities from the twenty-four TE volumes of ^23^Na-MRI acquisitions for signal calibration procedure. Linear fitting of ^23^Na signal decay from tubes ROI (Matlab) provided the slope (a) and intercept (b), which is used for calibration purposes (see below).

For each ROI (for details on ROI definition see section 2.6), we fitted the mean signal intensity across each of the 24 TEs with a biexponential model using the equation:

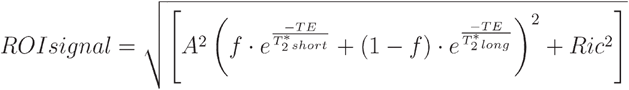

where *A* is an amplitude scaling term, *f* is the sodium signal fraction of the short T_2_* decay component and *Ric* refers to a Rician noise-related scaling parameter (Ridley et al., 2017). From the model we estimated a sodium signal fraction for short (*f*) and long (1-*f*) T_2_* decay. Note that these short and long fractions of the sodium signal are only present when the Na^+^ ions are present in an organized non-isotropic molecular environment. Indeed, in these environments, the electric quadrupolar moment of the sodium nucleus interacts strongly with the surrounding electric field gradient leading to a residual quadrupolar interaction responsible for the bi-exponential T_2_ relaxation with a short T_2_* and a long T_2_* depending on the motional regime of the environment. In contrast, in an isotropic non-restricted environment such as the cerebrospinal fluid, there is no residual quadrupolar interaction and all energetic transitions are equal and lead to a monoexponential decay with only one (long) T_2_* (Burstein & Springer, 2019). We calculated magnetization (M0) corresponding to the signal fraction estimated by the model in terms of the intercepts of the signal fraction components of the model, obtaining M0_SF_ = A·*f* and M0_LF_ = A·(1-*f*). Then, Na_SF_ and Na_LF_ were calculated with raw M0 signal values and the linear fit estimated over the tube phantoms, i.e. slope (a) and intercept (b):

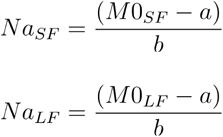

Finally, we calculated the total sodium concentration (TSC) in each ROI as follows:

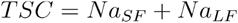

Each ROI had to be almost totally (> 99.9%) included in the brain mask of the individual patient and in the brain masks of at least half of all HC. Finally, to limit the partial volume effect related to CSF, patient ROIs with high CSF contains - estimated with structural image segmentation - relative to HC (|Z-score_CSF_| > 1.96) were also discarded. If after this final step a patient had no remaining EZ ROIs, the patient was entirely discarded from the analyzes, reducing the number of investigated patients from 22 to 19.

### 2.4. SEEG Recordings

Recordings were performed using intracerebral multiple contact electrodes (10–18 contacts with length 2 mm, diameter 0.8 mm, and 1.5 mm apart, Alcis, France). The electrodes were implanted using a stereotactic surgical robot ROSA™. Cranial CT scan was performed to verify the absence of any complication and the spatial accuracy of the implantation. CT/MRI data co-registration and 3D-reconstructions of patients’ brains with electrodes was performed using an in-house open-source software (EpiTools, (Medina Villalon et al., 2018)) to automatically localize the position of each electrode contact and display the results of signal analysis in each patient’s anatomy.

Signals were recorded on a 256-channel Natus system, sampled at 512 Hz and saved on a hard disk (16 bits/ sample) using no digital filter. Two hardware filters were present in the acquisition procedure: a high-pass filter (cut-off frequency equal to 0.16 Hz at -3 dB), and an anti-aliasing low-pass filter (cut-off frequency equal to 170 Hz at 512Hz).

### 2.5. SEEG-signal analysis

All signal analyzes were performed in a bipolar montage and computed using the open-source AnyWave software (Colombet et al., 2015) available at https://meg.univ-amu.fr/wiki/AnyWave. The epileptogenicity of different brain structures was assessed by quantitative SEEG-signal analysis using the Epileptogenicity Index (EI) (Bartolomei et al., 2008).^31^ The EI combines analysis of both spectral and temporal features of SEEG signals, respectively, related to the propensity of a brain area to generate fast discharges (12.4 – 127 Hz), and to how early this area becomes involved in seizure. A normalized EI value is used, ranging from 0 to 1. If there is no involvement of the brain structure, the EI = 0 (no epileptogenicity) whereas if the brain structure generates a rapid discharge and the time to seizure onset is minimal, the EI = 1 (maximal epileptogenicity).

In each patient, maximal EI values from at least three representative seizures were computed. We labeled each pair of bipolar SEEG contacts as belonging to the EZ, propagation zone (PZ) or non-involved zone (NIZ), as defined by EI, based on previous studies (Aubert et al., 2009; Lagarde et al., 2019). An EI value of 0.4 and higher was set as a threshold to define a structure as belonging to the EZ. The PZ was defined as brain areas with 0.1 < EI < 0.4, with sustained discharge during the seizure course. The NIZ was defined as all other brain structures.

### 2.6. ROI Definition

GARDEL software (Medina Villalon et al., 2018) provided the MRI voxel coordinates of electrode contacts allowing the definition of spherical ROIs for sodium and metabolite quantification in the patient’s native space. As SEEG recordings processed layout corresponds to the signal differential of two adjacent electrode contacts, a five mm radius spherical ROIs were positioned with a center between two adjacent electrode contacts (Ridley et al., 2017). ROIs corresponding to a poor SEEG signal quality - those located in white matter for instance - were discarded by expert (J.S.) inspection. To deal with possible contamination of the brain sodium signal by an over-representation of CSF in patients, we discarded ROIs with overly high Z-score_CSF_ (see section 2.3. MRI Data Processing). Anatomical references were the MPRAGE volumes for ^1^H-MRSI (3T) and the MP2RAGE volumes for ^23^Na-MRI (7T). ANTS brain extraction function was applied to all anatomical and ^23^Na-MRI images. Each subject’s (patients and controls) anatomical images were coregistered onto their respective ^1^H-MRSI and ^23^Na-MRI maps. Each individual anatomical volume was also spatially normalized onto the MNI 152 template to obtain the direct and reverse spatial transforms for each subject. ROIs were projected from the patient native space to MNI space, and back-projected from MNI to each HC native space (ANTS). This procedure allowed us to extract normative values of ^23^Na MRI and ^1^H-MRSI data from the control group for each ROI defined in each patient. Details about the numbers of ROIs for each category were summarized in supplementary Table 1.

### 2.7. Statistical Analysis

We performed a group comparison of sodium and metabolites levels across the three ROI classes, EZ, PZ and NIZ, to decipher subtle and specific homeostatic and metabolic modifications among patients compared to healthy controls. To account for brain sodium and metabolite variability across participants, sodium and metabolite levels had to be normalized across participants. Thus, each patient’s sodium concentrations and metabolites levels in each ROI were expressed as z-scores with respect to the same mean quantities from the corresponding ROI in healthy controls. Normalization of healthy controls’ sodium concentrations and metabolites levels was done using a leave-one-out procedure, z-scoring control’s quantities relative to the same ROI in other controls’ sodium concentrations and metabolites levels.

We compared patients and healthy controls, looking for different sodium concentration and/or metabolic change profiles between them, and between different region categories (described in ‘SEEG-signal analysis’). Significant group differences of z-scores were analyzed using a bootstrap two-tailed Welch’s t-test procedure (Efron & Tibshirani, 1993). Achieved significance levels (ASL) (equivalent to exact p-values) were obtained after one million random samplings, then corrected for multiple comparisons using false-discovery rate (FDR) (Benjamini & Hochberg, 1995).

In order to investigate the relationship between sodium concentrations and metabolites levels, we evaluated how well sodium concentration can be predicted from metabolite profiles. Hence, we performed a multiple linear regression analysis (Jobson, 1991) using Statsmodels (Seabold & Perktold, 2010) on all ROIs, with NAA, Cho and tCr z-scores as predictors to compute linear model for each ^23^Na-MRI measures (*f* and TSC) z-scores.

## 3. Results

### 3.1. Clinical features

Patients’ clinical characteristics are summarized in Table 1. Mean age at epilepsy onset was 13.2 years (range 0.1-40), mean duration of epilepsy was 18 years (range 4-31). Mean seizure frequency was 26 per month (range 2-120). Anatomical MRI was normal in fifteen (71%) and showed a structural abnormality in four (29%) cases. Thirteen out of twenty-one patients underwent curative surgical procedure following SEEG exploration. From the remaining eight, six patients were recused from surgery because of bilaterality of the epileptogenic zone or of a high risk of post-surgical functional impairment, one became seizure-free after SEEG-guided thermocoagulations and one refused surgery. The post-surgical outcome was favorable in eleven (Engel class I (seizure-free) and class II (almost seizure-free), n=11, 85%) and with worthwhile improvement in two cases (Engel class III, 15%). Epilepsy etiology, according to histopathological findings, was: focal cortical dysplasia (FCD) type I (non-detectable on MRI) in six patients, FCD type II in two, and slight gliosis in two. One patient underwent laser-guided interstitial thermosurgery for an MRI-diagnosed DNET, with no histology available.

From the initial sample of nineteen patients and according to the quality thresholds previously described, data analyzes were conducted in fifteen patients for ^23^Na-MRI and seventeen patients for ^1^H-MRSI (Table 1). Patients and healthy controls (HC) did not significantly differ in terms of age and sex, neither in the ^23^Na-MRI associated patient group (Wilcoxon rank sum, w = -0.36, p = 0.72; χ^2^ (1, N=33) = 0.26, p = 0.88) nor ^1^H-MRSI associated patient group (Wilcoxon rank sum, w = 0.58, p = 0.56; χ^2^ (1, N=42) = 0.038, p = 0.98).

### 3.2. Ion homeostasis and metabolic profiles in patients

TSC was increased in epileptic patients relative to corresponding ROIs in healthy controls in all types of electrophysiologically defined regions, (i.e. EZ, PZ and NIZ) with no significant differences between these three region categories within patients (see Table 2 and Figure 3.A). The short fraction *f* was significantly increased in EZ relative to corresponding ROIs in healthy controls, but was not significantly different compared to healthy controls in regions corresponding to PZ and NIZ. In patients, *f* within EZ was also significantly increased relative to PZ and to NIZ.

**Table 2.** Bootstrapped t-value followed by p (or ASL, the bootstrapped achieved significance level).

|  | EZ Vs HC | PZ Vs HC | NIZ Vs HC | EZ Vs PZ | EZ Vs NIZ | PZ Vs NIZ |
| --- | --- | --- | --- | --- | --- | --- |
| **f** | 2.96, $3.8 \cdot 10^{-3}$ | - | - | 2.51, 0.012 | 2.43, 0.015 | - |
| **TSC** | 3.73, $2.5 \cdot 10^{-4}$ | 5.35, $1.0 \cdot 10^{-6}$ | 12.83, $p < 1.0 \cdot 10^{-6}$ | - | - | - |
| **NAA** | -6.03, $p < 1.0 \cdot 10^{-6}$ | -4.51, $8.0 \cdot 10^{-6}$ | -9.46, $p < 1.0 \cdot 10^{-6}$ | - | -2.32, 0.022 | - |
| **Cho** | 3.31, $1.8 \cdot 10^{-3}$ | - | -2.75, $6.4 \cdot 10^{-3}$ | - | 3.94, $1.3 \cdot 10^{-4}$ | 2.47, 0.015 |
| **tCr** | - | - | -7.42, $p < 1.0 \cdot 10^{-6}$ | - | - | - |

FDR-correction yielded $p < 0.016$ as threshold for tested $^{23}\text{Na}$ -MRI measurements, and $p < 0.022$ for tested $^1\text{H}$ -MRSI.

**Figure 3:**
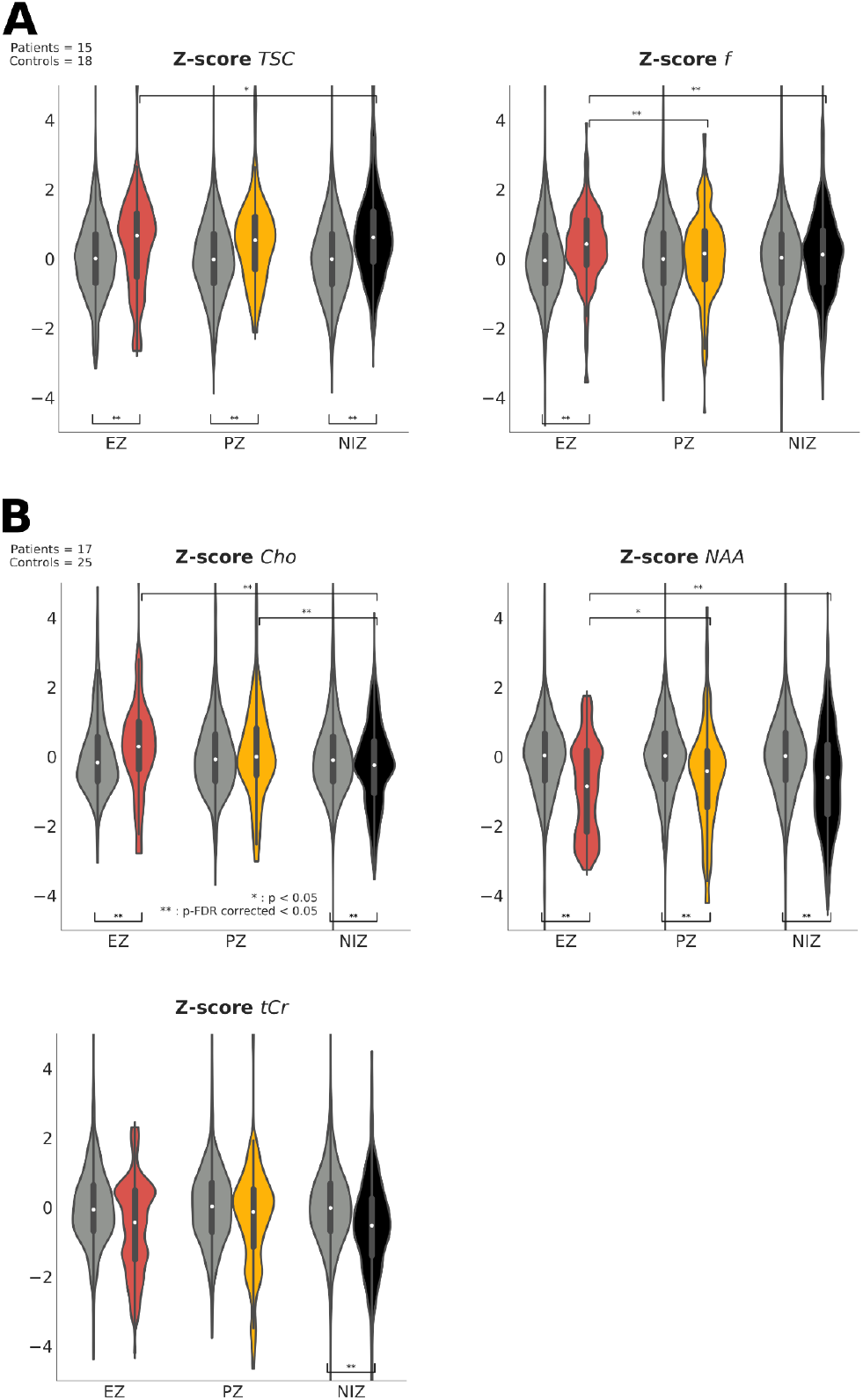
Violin plot of mean Z-scores of sodium (**A**) and metabolite estimates (**B**) observed in patients within epileptogenic zones (EZ, red), propagation zones (PZ, yellow) and non-involved zones (NIZ, black), compared to healthy controls (HC, gray). Asterisks indicate significant differences between patients and controls when under violins, and between regions within patients when above violins. *: p-uncorrected < 0.05. **: p-FDR < 0.05

In patients, NAA levels were significantly decreased in all types of regions relative to controls (see Table 2 and Figure 3.B). In addition, NAA was significantly decreased in EZ compared to PZ and NIZ. In patients relative to controls, tCho levels were significantly increased in EZ, decreased in NIZ and not significantly different in PZ; tCr levels were not significantly different in EZ and PZ but significantly decreased in NIZ.

### 3.3. Association between ionic and metabolic parameters

To study the relationship between sodium concentrations and metabolite levels, a multiple linear regression analysis was conducted. We evaluated the prediction of each ^23^Na-MRI measure from all ^1^H-MRSI measures in each region category, namely EZ, PZ and NIZ, setting Bonferroni corrected p for coefficient t-test was set at 0.05/ 2*3*3 = 0.0028.

Significant regression equations were found for the model predicting TSC from metabolites predictors alone in PZ (F(3, 80) = 10.76, p < 0.0028) and in NIZ (F(3, 283) = 16.25, p < 0.0028) but not in EZ (F(3, 34) = 1.29, p = 0.295). As mentioned previously, the significant regressions only partially thinly explained variations in TSC, with R² of .29 in PZ and a R² of .15 in NIZ. We observed a negative association between TSC and NAA in PZ (β = -0.87, p < 0.0028) and in NIZ (β = -0.45, p < 0.0028). We also found a negative association between TSC and Cho and a trend towards a positive association with tCr in NIZ (Table 3). Multiple linear regression analysis of *f* does not provide significant results in any ROI.

**Table 3.** Summary of multiple linear regression analysis for TSC and f.

|  |  | Parameters | Coefficients | p |  |  |
| --- | --- | --- | --- | --- | --- | --- |
| **TSC** | **EZ** | **Const** | 0,43 | 0,191 | Df Residuals | 34 |
|  |  | **Cho** | -0,05 | 0,837 | Df Model | 3 |
|  |  | **NAA** | -0,59 | 0,0743 | R<sup>2</sup> | 0.1 |
|  |  | **tCr** | 0,39 | 0,229 | F (p) | 1.29 (0.295) |
| | **PZ** | **Const** | 0,86 | $8,89 \cdot 10^{-5}$ | Df Residuals | 80 |
|  |  | **Cho** | -0,33 | 0,0787 | Df Model | 3 |
|  |  | **NAA** | -0,87 | **$9,02 \cdot 10^{-4*}$** | R<sup>2</sup> | 0.29 |
|  |  | **tCr** | 0,56 | **0,0408** | F (p) | **10.76 ($5.13 \cdot 10^{-6}$)** |
| | **NIZ** | **Const** | 0,94 | $1,60 \cdot 10^{-17}$ | Df Residuals | 283 |
|  |  | **Cho** | -0,29 | **$6,53 \cdot 10^{-3*}$** | Df Model | 3 |
|  |  | **NAA** | -0,45 | **$1,90 \cdot 10^{-6*}$** | R<sup>2</sup> | 0.15 |
|  |  | **tCr** | 0,25 | **0,0392** | F (p) | **16.25 ($8.96 \cdot 10^{-10}$)** |
| **f** | **EZ** | **Const** | 0,51 | $6,15 \cdot 10^{-3}$ | Df Residuals | 34 |
|  |  | **Cho** | -0,33 | **0,0114** | Df Model | 3 |
|  |  | **NAA** | 0,01 | 0,967 | R<sup>2</sup> | 0.18 |
|  |  | **tCr** | 0,24 | 0,17 | F (p) | 2.48 (0.0781) |
|  | **PZ** | **Const** | -0,12 | 0,448 | Df Residuals | 80 |
|  |  | **Cho** | 0,03 | 0,832 | Df Model | 3 |
|  |  | **NAA** | -0,28 | 0,154 | R<sup>2</sup> | 0.03 |
|  |  | **tCr** | 0,12 | 0,572 | F (p) | 0.94 (0.425) |
|  | **NIZ** | **Const** | 0,02 | 0,791 | Df Residuals | 283 |
|  |  | **Cho** | 0,11 | 0,158 | Df Model | 3 |
|  |  | **NAA** | 0,04 | 0,587 | R<sup>2</sup> | 0.01 |
|  |  | **tCr** | -0,16 | 0,0615 | F (p) | 1.4 (0.244) |

Bonferroni corrected p-values are highlighted in bold followed by an asterisk, while uncorrected p-values are in bold without asterisk.

## 4. Discussion

This work aimed to explore both *in vivo* ion homeostasis and metabolic alterations in focal drug-resistant epilepsy investigated by SEEG. We observed global brain impact in patients reflected by consistent multimodal alterations including an increase of TSC as well as decreases in NAA levels within all categories of electrically defined regions, namely EZ, PZ and NIZ. Interestingly, the EZ showed a characteristic pattern with significantly higher *f*, as well as significantly lower NAA and higher Cho levels. TSC exhibits strong association with metabolites, especially in the PZ and NIZ. These imaging features likely reflect both hyperexcitability and tissue alterations with a specific pattern of *f* for the epileptogenic tissues.

### 4.1. Deciphering sodium homeostasis processes with 7T ^23^Na MRI

Accumulation of TSC is a feature described in a number of neurological diseases including multiple sclerosis (Maarouf et al., 2017; Zaaraoui et al., 2012), amyotrophic lateral sclerosis (Grapperon et al., 2019), Hungtington’s disease (Reetz et al., 2012) and epilepsy (Ridley et al., 2017). Attempts to model processes involved in TSC increases have been proposed considering *in vitro* experiments showing increases of intracellular sodium concentrations in multiple sclerosis (Waxman, 2006). In this model, an increase in TSC – the total sodium concentration - is usually associated with an elevation of *intracellular* sodium concentration due to an influx of sodium resulting from a dysfunction of the sodium potassium pump (Na+/K+ pump) (Pike et al., 1985). However, though sensitive, TSC is not specific to altered homeostasis between sodium compartments.

TSC alterations in humans *in vivo* have largely been demonstrated at 3T. Here, to improve our understanding of the dysregulation mechanisms affecting the sodium ion homeostasis in epilepsy, we used 7T ^23^Na MRI. Ultra-high field offers the opportunity to study the complexity of the T_2_^*^ decays of the ^23^Na MR signal influenced by the quadrupolar relaxation. Indeed, multi-exponential relaxations of this three-half spin reflect the time-dependent relative position of the quadrupolar moments of sodium nuclei and the electric field gradients of charged molecules at their vicinity. The multi-TE density adapted radial ^23^Na, ultra-short echo time (UTE) approach used here permits the characterization of the signal decay components through a bi-exponential fit. While related, the signal fraction of the short T_2_* decay component (*f*), and the total sodium concentration (TSC) may reflect different consequences of ion homeostasis dysregulation (Ridley et al., 2018).

Normal physiological neuronal activity has been observed to dynamically alter sodium signal across different TEs, in a manner consilient with fMRI and consistent with physiological mechanisms that should dominate at different points of ‘activation’ (Bydder et al., 2019) Using dynamic sodium MRI acquisition compared to BOLD functional MRI, variations in sodium signals recorded at different TE (0.2ms, 10ms, 19ms) during a right finger tapping task showed a slightly increased TSC (TE=0.2ms) in the activated left contralateral motor area interpreted as increased cerebral blood volume, and a more drastic signal decrease at longer TEs considered, at least in part, to a decrease in extracellular contributions due a reduced extracellular volume fraction (Antonio et al., 2016; Dietzel et al., 1982; Lux et al., 1986), while reverse signal variations were observed in the deactivated motor area ipsilateral to movement. The *f* metric, reflecting the variations in the ratio of apparent short and long T_2_* sodium signal decays, enables to pool in a single parameter these effects seen at different TEs, and has the potential to locate regions with abnormal excitability as shown in the present study.

### 4.2. Sodium changes in epilepsy

In the context of epilepsy, abnormal TSC increase has been found in EZ and to a lesser extent also in other regions of the brain at 3T (Ridley et al., 2017). In addition, rat models of acquired epilepsy have reported persistent TSC increases in affected cortices in response to kainate-induced epileptogenesis (Mori et al., 2000; Y. Wang et al., 1996). TSC may incorporate various processes including both structural reorganization and dysregulation of ionic homeostasis. Indeed, several structural modifications can impact ^23^Na-MRI signals, particularly changes in the extracellular volume (ECV). Cell shrinkage and cerebral atrophy could lead to ECV increase, and subsequently to increases in TSC. Both have been reported in epilepsy, localized in the epileptic areas, but also often extended to the non-epileptogenic regions (Bernhardt et al., 2009; Dingledine et al., 2014; Liu et al., 2005; Voets et al., 2017). Thus, even though excessively high CSF levels (Z-score_CSF_ > 1.95) led to the removal of a ROI from consideration, its contribution cannot be entirely ruled out. In addition, ECV increase was recently shown to be related to neuronal excitability (Colbourn et al., 2019). Moreover, increases of perivascular space were also reported in epilepsy (Feldman et al., 2018, 2019), and could contribute to TSC accumulation due to the CSF surrounding the vessels. Recently, venous blood was also demonstrated to have an impact on total sodium signal (Driver et al., 2020), leading to overestimation of sodium concentration measures in case of atrophy. In addition, reactive astrogliosis and microgliosis (Devinsky et al., 2013; Seifert & Steinhäuser, 2013; Sofroniew & Vinters, 2010) can be associated to epilepsy implying changes of astrocytes proportion, size (Boscia et al., 2016) and ion homeostasis, leading to surrounding cell homeostasis disruption (Karus et al., 2015). Interestingly, TSC was also shown to correlate with conductivity at 3T (Liao et al., 2019).

Beyond general explanations for alterations of TSC in epilepsy, cortices subject to different epileptiform manifestations (EZ, PZ; NIZ) are likely to differ in underlying pathological mechanisms, something that was probed in the current work through the use of a multiparametric approach. While keeping in mind that both sodium signal fractions (i.e. short and long) will contain contributions from extracellular sodium – albeit with potentially different weightings – a plausible reason for the concomitant EZ-specific increase relative to both HC and other cortices of both *f* and TSC is an increase of intracellular sodium concentration. This would be consistent with a range of known mechanisms associated with the EZ while TSC increase outside the EZ in the absence of changes in f may could be related to structural changes combined with preserved perfusion, which is usually decreased in the EZ during the interictal period (Kojan et al., 2021; Y.-H. Wang et al., 2018).

Sodium homeostasis dysregulation in the EZ could be induced by several mechanisms affecting ion channels such as (i) alterations in type II and III voltage gated sodium channel (VGNC) properties (Bartolomei et al., 1997; Gastaldi et al., 1997; Gorter et al., 2010; Lombardo et al., 1996), (ii) incomplete inactivation of sodium channels and a consequent increase in persistent sodium currents (Mantegazza et al., 2010; Oliva et al., 2012) and (iii) the reduced efficiency of clearance by the Na+/K+ pump induced by a lack in ATP supplies (Folbergrová & Kunz, 2012; Grisar et al., 1992; Kovac et al., 2017).

Other mechanisms secondary to hyperexcitability or energy failure affecting astrocytes could also lead to an alteration of sodium homeostasis and an increase in intracellular sodium (Gerkau et al., 2017; Kirischuk et al., 2012; Rose & Karus, 2013). Indeed, as a consequence of hyperexcitability, astrocytes may uptake sodium through various tranporters(sodium-potassium-chloride co-transporter (NKCC1), sodium-bicarbonate co-transporter (NBC), sodium-proton exchanger (NHE) and sodium-calcium exchanger (NCX)) increasing the intracellular sodium concentrations. Heightened intracellular sodium concentrations reduce the Electrochemical gradient for glutamate uptake (Karus et al., 2015) by the excitatory amino acid transporter (EAAT), a transmembrane transporter of sodium and glutamate from peri-synaptic extracellular space into astrocytes, ensuring ion and neurotransmitter clearance. Then extracellular glutamate concentration increases and eventually downregulates EAAT (Rose & Karus, 2013); as astrocytes store energy supplies, astrocytic energy failure is critical for ion homeostasis of both astrocytes and the surrounding neurons. The reduced availability of ATP associated with increases in intracellular sodium also leads to the dysregulation of homeostasis for other ions such as potassium, calcium and proton, resulting in excitotoxicity (Gerkau et al., 2017). During hyperexcitability, intracellular pH decreases which leads to sodium uptake via NBC, VGNC or Na+/K+ pump among others. This reduced intracellular pH was suggested to result from lack of NHE1 (Zhao et al., 2016). The relationship between reduced intracellular pH and increased intracellular sodium was also shown in epilepsy during hepatic encephalitis (Kelly et al., 2009) with ammonium intoxication. The intoxication of cerebral tissue promotes intracellular pH increase (in particular in astrocytes) resulting in intracellular sodium increase.

### 4.3. Metabolic changes

The multimodal nature of our investigation provides further evidence of an alteration in ionic homeostasis in epilepsy due to a disruption of metabolic energy supply. The decreased NAA we observed has been consistently associated with neuronal death, mitochondrial dysfunction (Stefano et al., 1995) and subsequent ATP decrease (Vagnozzi et al., 2007). NAA decrease in the EZ has been widely reported since the 90’s (Guye et al., 2005; Hugg et al., 1993; Kuzniecky et al., 1998; Petroff et al., 2003; Simister et al., 2002; van der Hel et al., 2013). While more recent research indicates that decreases in NAA extends to other regions, those involved in seizures remain the most affected (Guye et al., 2005; Lundbom et al., 2001; Mueller et al., 2011) as is the case for TSC as well as NAA in this study. This finding is in line with our energetic failure hypothesis, however, it should be noted we were unable to observe a significant association between these two measures when explored by multiple linear regression.

Increased Cho has also been reported in temporal lobe epilepsy (Achten et al., 1997; Simister et al., 2009) and is frequently associated with cellular proliferation and increased membrane turnover (Miller, 1991; Urenjak et al., 1993). These processes are likely to be linked to structural changes associated with epileptogenic lesions such as tumors, tumor-like tissue and gliosis (van der Hel et al., 2013). The Cho decrease in NIZ was rather unexpected and has not to our knowledge been previously reported. It could result from reduced cellular density or neuronal loss extending beyond the EZ.

In our cohort, tCr was significantly decreased in NIZ only. Heighted tCr was previously related to decreased intracellular energy status or reactive astrocytes in the litterature (Achten, 1998; Urenjak et al., 1993). This result also points to alterations beyond EZ potentially accompanying structural changes and cognitive co-morbidities (Kreis et al., 1992; Kreis & Ross, 1992).

### 4.4. Technical limitations

For both ^23^Na-MRI and ^1^H-MRSI, CSF signal contribution is critical as it can clearly bias the data. MIDAS software handled this bias for ^1^H-MRSI data (Lecocq et al., 2015). For ^23^Na-MRI we corrected for partial volume effect and removed ROIs with remaining CSF after segmentation. We designed a quality check procedure inspired by (Ridley et al., 2017) in order to get rid of this CSF contribution. Despite this, it is impossible to totally delimit the partial volume effect completely for the moment, neither for ^1^H-MRSI nor ^23^Na-MRI. Another limitation is inherent to the SEEG procedure, which suffers from limited spatial sampling, whereas MRI gives access to information across the entire brain. Conversely, the pathological specificity offered by SEEG is considered gold-standard and beyond what is possible with anatomically defined and atlas derived ROIs.

### 4.5. Conclusion and Perspectives

An increase of the signal fraction of the short T_2_* decay component (*f*) was found to be associated with the EZ whereas increased TSC was not limited to epileptogenic regions. Taken together with the patterns of metabolite changes our results are in line with the energetic failure hypothesis in epileptic regions associated with widespread tissue changes beyond electrically abnormal areas. Here, a multimodal approach has allowed parallel insights supporting pathological processes in epilepsy. This and additional combinations - including for example ^31^P-MRSI, which provides information about energy metabolism via ATP quantification and pH estimation - could further strengthen the delineation of the EZ as this *non-invasive* information would complement the current presurgical evaluation in patients suffering from drug resistant focal epilepsy.

## Supporting information

Supplemental Figure 1 : Panel of spectra as explored during quality assessment.

Supplemental Table 1. Total number of ROIs per zone

Supplemental Table 2. ROIs 23Na-MRI and 1H-MRSI measures

## Data Availability

The data that support the findings of this study are available on request from the
corresponding author. The data are not publicly available due to sensitive information that
could compromise the privacy of research participants.

## Abbreviations

AC-PC: Anterior Commissure - Posterior Commissure
ASL: Achieved Significance Level
Cho: Choline Compounds
CRLB: Cramer-Rao Lower Bound
DNET: Dysembryoplastic Neuroepithelial Tumor
EAAT: Glutamate Transporter
ECV: Extracellular Volume
EI: Epileptogenicity Index
EZ: Epileptogenic Zones
f: fraction of sodium signal with short T_2_* decays
FCD: Focal Cortical Dysplasia
FDR: False Discovery Rate
^18^F-FDG-PET: Fluorodeoxyglucose PET
FOV: Field of View
GRAPPA: GeneRalized Autocalibrating Partial Parallel Acquisition
^1^H: Hydrogen
HC: Healthy Controls
MIDAS: Metabolite Imaging and Data Analyses System
MNI: Montreal Neurological Institute
MP2RAGE: Magnetization Prepared 2 Rapid Acquisition Gradient Echoes
MPRAGE: Magnetization Prepared Rapid Acquisition Gradient Echoes
EPSI: Echo Planar Spectroscopic Imaging
^23^Na: Sodium
NAA: N-Acetyl Aspartate
Na_LF_: apparent concentrations of sodium with long T_2_* decays
Na_SF_: apparent concentrations of sodium with short apparent T_2_* decay
NBC: Bicarbonate Sodium Cotransporter
NCX: Sodium Calcium Exchanger
NHE: Sodium Hydrogen Antiporter
NIZ: Non Involved Zones
Na+/K+ pump: Sodium Potassium pump
NKCC1: Sodium Potassium Chloride cotransporter
pH: Potential of Hydrogen
PZ: Propagation Zones
QED: Quality Electrodynamics
ROI: Region of Interest
SEEG: Stereotactic EEG
T_2_*_long_: Sodium longT_2_* relaxation time
T_2_*_short_: Sodium short T_2_* relaxation time
tCr: Total Creatine
TE: Echo Time
TI: Inversion Time
TR: Repetition Time
TSC: Total Sodium Concentration
UTE: Ultrashort Echo Time
VGNC: Voltage Gated Sodium Channels

## Acknowledgements

The authors would like to thank L. Pini, C. Costes, and V. Gimenez for data acquisition and study logistics. We would also like to thank A. Ivanov and C. Bernard for helpful discussions.

## Data Availability Statement

The data that support the findings of this study are available on request from the corresponding author. The data are not publicly available due to sensitive information that could compromise the privacy of research participants.

## CRediT authorship contribution statement

**Mikhael Azilinon**: Investigation, Methodology, Data Curation, Formal Analysis, Writing - original draft. **Julia Scholly**: Resources, Data Curation, Formal Analysis, Writing - review and editing. **Wafaa Zaaraoui**: Funding Acquisition, Writing - review and editing. **Samuel Medina Villalon:** Methodology. **Patrick Viout:** Formal Analysis. **Tangi Roussel:** Methodology, Writing - review and editing. **Mohamed Mounir El Mendili:** Methodology, Writing - review and editing. **Ben Ridley:** Methodology, Writing - review and editing. **Jean-Philippe Ranjeva:** Supervision, Validation, Methodology, Writing - original draft. **Fabrice Bartolomei:** Funding Acquisition, Resources, Writing - review and editing. **Viktor Jirsa:** Funding Acquisition, Supervision, Writing - review and editing. **Maxime Guye:** Project Administration, Funding Acquisition, Resources, Conceptualization, Supervision, Validation, Writing - original draft.

## References

Achten, E. (1998). Aspects of proton MR spectroscopy in the seizure patient. Neuroimaging Clinics of North America, 8(4), 849–862.

Achten, E., Boon, P., Kerckhove, T. V. D., Caemaert, J., Reuck, J. D., & Kunnen, M. (1997). Value of Single-Voxel Proton MR Spectroscopy in Temporal Lobe Epilepsy. 9.

Aja-Fernandez, S., Alberola-Lopez, C., & Westin, C. (2008). Noise and Signal Estimation in Magnitude MRI and Rician Distributed Images: A LMMSE Approach. IEEE Transactions on Image Processing, 17(8), 1383–1398. https://doi.org/10.1109/TIP.2008.925382

Antonio, L. L., Anderson, M. L., Angamo, E. A., Gabriel, S., Klaft, Z.-J., Liotta, A., Salar, S., Sandow, N., & Heinemann, U. (2016). In vitro seizure like events and changes in ionic concentration. Journal of Neuroscience Methods, 260, 33–44. https://doi.org/10.1016/j.jneumeth.2015.08.014

Aubert, S., Wendling, F., Regis, J., McGonigal, A., Figarella-Branger, D., Peragut, J.-C., Girard, N., Chauvel, P., & Bartolomei, F. (2009). Local and remote epileptogenicity in focal cortical dysplasias and neurodevelopmental tumours. Brain, 132(11), 3072–3086. https://doi.org/10.1093/brain/awp242

Avants, B. B., Tustison, N. J., Song, G., Cook, P. A., Klein, A., & Gee, J. C. (2011). A reproducible evaluation of ANTs similarity metric performance in brain image registration. NeuroImage, 54(3), 2033–2044. https://doi.org/10.1016/j.neuroimage.2010.09.025

Bartolomei, F., Chauvel, P., & Wendling, F. (2008). Epileptogenicity of brain structures in human temporal lobe epilepsy: A quantified study from intracerebral EEG. Brain, 131(7), 1818–1830. https://doi.org/10.1093/brain/awn111

Bartolomei, F., Gastaldi, M., Massacrier, A., Planells, R., Nicolas, S., & Cau, P. (1997). Changes in the mRNAs encoding subtypes I, II and III sodium channel alpha subunits following kainate-induced seizures in rat brain. Journal of Neurocytology, 26(10), 667–678. https://doi.org/10.1023/a:1018549928277

Benjamini, Y., & Hochberg, Y. (1995). Controlling the False Discovery Rate: A Practical and Powerful Approach to Multiple Testing. Journal of the Royal Statistical Society. Series B (Methodological), 57(1), 289–300.

Bernhardt, B. C., Rozen, D. A., Worsley, K. J., Evans, A. C., Bernasconi, N., & Bernasconi, A. (2009). Thalamo–cortical network pathology in idiopathic generalized epilepsy: Insights from MRI-based morphometric correlation analysis. NeuroImage, 46(2), 373–381. https://doi.org/10.1016/j.neuroimage.2009.01.055

Boscia, F., Begum, G., Pignataro, G., Sirabella, R., Cuomo, O., Casamassa, A., Sun, D., & Annunziato, L. (2016). Glial Na ^+^-dependent ion transporters in pathophysiological conditions: Glial Na ^+^-dependent Transporters in CNS Diseases. Glia, 64(10), 1677–1697. https://doi.org/10.1002/glia.23030

Burstein, D., & Springer, C. S. (2019). Sodium MRI revisited. Magnetic Resonance in Medicine, 82(2), 521–524. https://doi.org/10.1002/mrm.27738

Bydder, M., Zaaraoui, W., Ridley, B., Soubrier, M., Bertinetti, M., Confort-Gouny, S., Schad, L., Guye, M., & Ranjeva, J.-P. (2019). Dynamic 23Na MRI -A non-invasive window on neuroglial-vascular mechanisms underlying brain function. NeuroImage, 184, 771–780. https://doi.org/10.1016/j.neuroimage.2018.09.071

Colbourn, R., Naik, A., & Hrabetova, S. (2019). ECS Dynamism and Its Influence on Neuronal Excitability and Seizures. Neurochemical Research, 44(5), 1020–1036. https://doi.org/10.1007/s11064-019-02773-w

Colombet, B., Woodman, M., Badier, J. M., & Bénar, C. G. (2015). AnyWave: A cross-platform and modular software for visualizing and processing electrophysiological signals. Journal of Neuroscience Methods, 242, 118–126. https://doi.org/10.1016/j.jneumeth.2015.01.017

Devinsky, O., Vezzani, A., Najjar, S., De Lanerolle, N. C., & Rogawski, M. A. (2013). Glia and epilepsy: Excitability and inflammation. Trends in Neurosciences, 36(3), 174–184. https://doi.org/10.1016/j.tins.2012.11.008

Dietzel, I., Heinemann, U., Hofmeier, G., & Lux, H. D. (1982). Stimulus-induced changes in extracellular Na+ and Cl-concentration in relation to changes in the size of the extracellular space. Experimental Brain Research, 46(1), 73–84. https://doi.org/10.1007/BF00238100

Dingledine, R., Varvel, N. H., & Dudek, F. E. (2014). When and How Do Seizures Kill Neurons, and Is Cell Death Relevant to Epileptogenesis? In H. E. Scharfman & P. S. Buckmaster (Eds.), Issues in Clinical Epileptology: A View from the Bench (Vol. 813, pp. 109–122). Springer Netherlands. https://doi.org/10.1007/978-94-017-8914-1_9

Donadieu, M., Le Fur, Y., Maarouf, A., Gherib, S., Ridley, B., Pini, L., Rapacchi, S., Confort-Gouny, S., Guye, M., Schad, L. R., Maudsley, A. A., Pelletier, J., Audoin, B., Zaaraoui, W., & Ranjeva, J.-P. (2019). Metabolic counterparts of sodium accumulation in multiple sclerosis: A whole brain 23Na-MRI and fast 1H-MRSI study. Multiple Sclerosis Journal, 25(1), 39–47. https://doi.org/10.1177/1352458517736146

Driver, I. D., Stobbe, R. W., Wise, R. G., & Beaulieu, C. (2020). Venous contribution to sodium MRI in the human brain. Magnetic Resonance in Medicine, 83(4), 1331–1338. https://doi.org/10.1002/mrm.27996

Efron, B., & Tibshirani, R. J. (1993). An Introduction to the Bootstrap. Springer US. https://doi.org/10.1007/978-1-4899-4541-9

Feldman, R. E., Delman, B. N., Pawha, P. S., Dyvorne, H., Rutland, J. W., Yoo, J., Fields, M. C., Marcuse, L. V., & Balchandani, P. (2019). 7T MRI in epilepsy patients with previously normal clinical MRI exams compared against healthy controls. PLOS ONE, 14(3), e0213642. https://doi.org/10.1371/journal.pone.0213642

Feldman, R. E., Rutland, J. W., Fields, M. C., Marcuse, L. V., Pawha, P. S., Delman, B. N., & Balchandani, P. (2018). Quantification of perivascular spaces at 7 T: A potential MRI biomarker for epilepsy. Seizure, 54, 11–18. https://doi.org/10.1016/j.seizure.2017.11.004

Folbergrová, J., & Kunz, W. S. (2012). Mitochondrial dysfunction in epilepsy. Mitochondrion, 12(1), 35–40. https://doi.org/10.1016/j.mito.2011.04.004

Gastaldi, M., Bartolomei, F., Massacrier, A., Planells, R., Robaglia-Schlupp, A., & Cau, P. (1997). Increase in mRNAs encoding neonatal II and III sodium channel a-isoforms during kainate-induced seizures in adult rat hippocampus. 12.

Gerkau, N. J., Rakers, C., Petzold, G. C., & Rose, C. R. (2017). Differential effects of energy deprivation on intracellular sodium homeostasis in neurons and astrocytes: Effects of energy deprivation on cellular sodium. Journal of Neuroscience Research, 95(11), 2275–2285. https://doi.org/10.1002/jnr.23995

Gorter, J. A., Zurolo, E., Iyer, A., Fluiter, K., Van Vliet, E. A., Baayen, J. C., & Aronica, E. (2010). Induction of sodium channel Nax (SCN7A) expression in rat and human hippocampus in temporal lobe epilepsy: SCN7A Expression in TLE. Epilepsia, 51(9), 1791–1800. https://doi.org/10.1111/j.1528-1167.2010.02678.x

Grapperon, A.-M., Ridley, B., Verschueren, A., Maarouf, A., Confort-Gouny, S., Fortanier, E., Schad, L., Guye, M., Ranjeva, J.-P., Attarian, S., & Zaaraoui, W. (2019). Quantitative Brain Sodium MRI Depicts Corticospinal Impairment in Amyotrophic Lateral Sclerosis. Radiology, 292(2), 422–428. https://doi.org/10.1148/radiol.2019182276

Grimaldi, S., El Mendili, M. M., Zaaraoui, W., Ranjeva, J.-P., Azulay, J.-P., Eusebio, A., & Guye, M. (2021). Increased Sodium Concentration in Substantia Nigra in Early Parkinson’s Disease: A Preliminary Study With Ultra-High Field (7T) MRI. Frontiers in Neurology, 12, 1610. https://doi.org/10.3389/fneur.2021.715618

Grisar, T., Guillaume, D., & Delgado-Escuet, A. V. (1992). Contribution of Na+,K+-ATPase to focal epilepsy: A brief review. Epilepsy Research, 12(2), 141–149. https://doi.org/10.1016/0920-1211(92)90034-Q

Guye, M., Le Fur, Y., Confort-Gouny, S., Ranjeva, J.-P., Bartolomei, F., Régis, J., Raybaud, C. A., Chauvel, P., & Cozzone, P. J. (2002). Metabolic and Electrophysiological Alterations in Subtypes of Temporal Lobe Epilepsy: A Combined Proton Magnetic Resonance Spectroscopic Imaging and Depth Electrodes Study. Epilepsia, 43(10), 1197–1209. https://doi.org/10.1046/j.1528-1157.2002.05102.x

Guye, M., Ranjeva, J. P., Le Fur, Y., Bartolomei, F., Confort-Gouny, S., Regis, J., Chauvel, P., & Cozzone, P. J. (2005). 1H-MRS imaging in intractable frontal lobe epilepsies characterized by depth electrode recording. NeuroImage, 26(4), 1174–1183. https://doi.org/10.1016/j.neuroimage.2005.03.023

Hugg, J. W., Laxer, K. D., Matson, G. B., Maudsley, A. A., & Weiner, M. W. (1993). Neuron loss localizes human temporal lobe epilepsy by in vivo proton magnetic resonance spectroscopic imaging. Annals of Neurology, 34(6), 788–794. https://doi.org/10.1002/ana.410340606

Isnard, J., Taussig, D., Bartolomei, F., Bourdillon, P., Catenoix, H., Chassoux, F., Chipaux, M., Clémenceau, S., Colnat-Coulbois, S., Denuelle, M., Derrey, S., Devaux, B., Dorfmüller, G., Gilard, V., Guenot, M., Job-Chapron, A.-S., Landré, E., Lebas, A., Maillard, L., … Sauleau, P. (2018). French guidelines on stereoelectroencephalography (SEEG). Neurophysiologie Clinique, 48(1), 5–13. https://doi.org/10.1016/j.neucli.2017.11.005

Jobson, J. D. (1991). Multiple Linear Regression. In J. D. Jobson (Ed.), Applied Multivariate Data Analysis: Regression and Experimental Design (pp. 219–398). Springer. https://doi.org/10.1007/978-1-4612-0955-3_4

Karus, C., Mondragão, M. A., Ziemens, D., & Rose, C. R. (2015). Astrocytes restrict discharge duration and neuronal sodium loads during recurrent network activity: Astrocytes Restrict Neuronal Sodium Loads. Glia, 63(6), 936–957. https://doi.org/10.1002/glia.22793

Kelly, T., Kafitz, K. W., Roderigo, C., & Rose, C. R. (2009). Ammonium-evoked alterations in intracellular sodium and pH reduce glial glutamate transport activity. Glia, 57(9), 921–934. https://doi.org/10.1002/glia.20817

Kirischuk, S., Parpura, V., & Verkhratsky, A. (2012). Sodium dynamics: Another key to astroglial excitability? Trends in Neurosciences, 35(8), 497–506. https://doi.org/10.1016/j.tins.2012.04.003

Kojan, M., Gajdoš, M., Říha, P., Doležalová, I., Řehák, Z., & Rektor, I. (2021). Arterial Spin Labeling is a Useful MRI Method for Presurgical Evaluation in MRI-Negative Focal Epilepsy. Brain Topography, 34(4), 504–510. https://doi.org/10.1007/s10548-021-00833-5

Kovac, S., Dinkova Kostova, A., Herrmann, A., Melzer, N., Meuth, S., & Gorji, A. (2017). Metabolic and Homeostatic Changes in Seizures and Acquired Epilepsy—Mitochondria, Calcium Dynamics and Reactive Oxygen Species. International Journal of Molecular Sciences, 18(9), 1935. https://doi.org/10.3390/ijms18091935

Kreis, R., Ernst, T., & Ross, B. D. (1993). Development of the human brain: In vivo quantification of metabolite and water content with proton magnetic resonance spectroscopy. Magnetic Resonance in Medicine, 30(4), 424–437. https://doi.org/10.1002/mrm.1910300405

Kreis, R., & Ross, B. D. (1992). Cerebral metabolic disturbances in patients with subacute and chronic diabetes mellitus: Detection with proton MR spectroscopy. Radiology, 184(1), 123–130. https://doi.org/10.1148/radiology.184.1.1319074

Kreis, R., Ross, B. D., Farrow, N. A., & Ackerman, Z. (1992). Metabolic disorders of the brain in chronic hepatic encephalopathy detected with H-1 MR spectroscopy. Radiology. https://doi.org/10.1148/radiology.182.1.1345760

Kuzniecky, R., Hugg, J. W., Hetherington, H., Butterworth, E., Bilir, E., Faught, E., & Gilliam, F. (1998). Relative utility of 1H spectroscopic imaging and hippocampal volumetry in the lateralization of mesial temporal lobe epilepsy. Neurology, 51(1), 66–71. https://doi.org/10.1212/WNL.51.1.66

Lagarde, S., Scholly, J., Popa, I., Valenti-Hirsch, M. P., Trebuchon, A., McGonigal, A., Milh, M., Staack, A. M., Lannes, B., Lhermitte, B., Proust, F., Benmekhbi, M., Scavarda, D., Carron, R., Figarella-Branger, D., Hirsch, E., & Bartolomei, F. (2019). Can histologically normal epileptogenic zone share common electrophysiological phenotypes with focal cortical dysplasia? SEEG-based study in MRI-negative epileptic patients. Journal of Neurology, 266(8), 1907–1918. https://doi.org/10.1007/s00415-019-09339-4

Larivière, S., Bernasconi, A., Bernasconi, N., & Bernhardt, B. C. (2021). Connectome biomarkers of drug-resistant epilepsy. Epilepsia, 62(1), 6–24. https://doi.org/10.1111/epi.16753

Lecocq, A., Le Fur, Y., Maudsley, A. A., Le Troter, A., Sheriff, S., Sabati, M., Donnadieu, M., Confort-Gouny, S., Cozzone, P. J., Guye, M., & Ranjeva, J.-P. (2015). Whole-brain quantitative mapping of metabolites using short echo three-dimensional proton MRSI: 3D^-1^ H-MRSI Covering the Whole Brain. Journal of Magnetic Resonance Imaging, 42(2), 280–289. https://doi.org/10.1002/jmri.24809

Liao, Y., Lechea, N., Magill, A. W., Worthoff, W. A., Gras, V., & Shah, N. J. (2019). Correlation of quantitative conductivity mapping and total tissue sodium concentration at 3T/4T. Magnetic Resonance in Medicine, 82(4), 1518–1526. https://doi.org/10.1002/mrm.27787

Liu, R. S. N., Lemieux, L., Bell, G. S., Sisodiya, S. M., Bartlett, P. A., Shorvon, S. D., Sander, J. W. A. S., & Duncan, J. S. (2005). Cerebral Damage in Epilepsy: A Population-based Longitudinal Quantitative MRI Study. Epilepsia, 46(9), 1482–1494. https://doi.org/10.1111/j.1528-1167.2005.51603.x

Lombardo, A. J., Kuzniecky, R., Powers, R. E., & Brown, G. B. (1996). Altered brain sodium channel transcript levels in human epilepsy. Molecular Brain Research, 35(1–2), 84–90. https://doi.org/10.1016/0169-328X(95)00194-W

Lundbom, N., Gaily, E., Vuori, K., Paetau, R., Liukkonen, E., Rajapakse, J. C., Valanne, L., Häkkinen, A., & Granström, M. (2001). Proton Spectroscopic Imaging Shows Abnormalities in Glial and Neuronal Cell Pools in Frontal Lobe Epilepsy. Epilepsia, 42(12), 1507–1514. https://doi.org/10.1046/j.1528-1157.2001.15301.x

Lux, H. D., Heinemann, U., & Dietzel, I. (1986). Ionic changes and alterations in the size of the extracellular space during epileptic activity. Advances in Neurology, 44, 619–639.

Maarouf, A., Audoin, B., Pariollaud, F., Gherib, S., Rico, A., Soulier, E., Confort-Gouny, S., Guye, M., Schad, L., Pelletier, J., Ranjeva, J.-P., & Zaaraoui, W. (2017). Increased total sodium concentration in gray matter better explains cognition than atrophy in MS. Neurology, 88(3), 289–295. https://doi.org/10.1212/WNL.0000000000003511

Madelin, G., Lee, J.-S., Regatte, R. R., & Jerschow, A. (2014). Sodium MRI: Methods and applications. Progress in Nuclear Magnetic Resonance Spectroscopy, 79, 14–47. https://doi.org/10.1016/j.pnmrs.2014.02.001

Mantegazza, M., Curia, G., Biagini, G., Ragsdale, D. S., & Avoli, M. (2010). Voltage-gated sodium channels as therapeutic targets in epilepsy and other neurological disorders. The Lancet Neurology, 9(4), 413–424. https://doi.org/10.1016/S1474-4422(10)70059-4

Maudsley, A. A., Darkazanli, A., Alger, J. R., Hall, L. O., Schuff, N., Studholme, C., Yu, Y., Ebel, A., Frew, A., Goldgof, D., Gu, Y., Pagare, R., Rousseau, F., Sivasankaran, K., Soher, B. J., Weber, P., Young, K., & Zhu, X. (2006). Comprehensive processing, display and analysis for in vivo MR spectroscopic imaging. NMR in Biomedicine, 19(4), 492–503. https://doi.org/10.1002/nbm.1025

Medina Villalon, S., Paz, R., Roehri, N., Lagarde, S., Pizzo, F., Colombet, B., Bartolomei, F., Carron, R., & Bénar, C.-G. (2018). EpiTools, A software suite for presurgical brain mapping in epilepsy: Intracerebral EEG. Journal of Neuroscience Methods, 303, 7–15. https://doi.org/10.1016/j.jneumeth.2018.03.018

Miller, B. L. (1991). A review of chemical issues in1H NMR spectroscopy:N-acetyl-l-aspartate, creatine and choline. NMR in Biomedicine, 4(2), 47–52. https://doi.org/10.1002/nbm.1940040203

Moffett, J. R., Arun, P., Ariyannur, P. S., & Namboodiri, A. M. A. (2013). N-Acetylaspartate reductions in brain injury: Impact on post-injury neuroenergetics, lipid synthesis, and protein acetylation. Frontiers in Neuroenergetics, 5. https://doi.org/10.3389/fnene.2013.00011

Mori, Y., Kondziolka, D., Balzer, J., Fellows, W., Flickinger, J. C., Lunsford, L. D., & Thulborn, K. R. (2000). Effects of stereotactic radiosurgery on an animal model of hippocampal epilepsy. Neurosurgery, 46(1), 157–165; discussion 165-168.

Mueller, S. G., Ebel, A., Barakos, J., Scanlon, C., Cheong, I., Finlay, D., Garcia, P., Weiner, M. W., & Laxer, K. D. (2011). Widespread extrahippocampal NAA/(Cr+Cho) abnormalities in TLE with and without mesial temporal sclerosis. Journal of Neurology, 258(4), 603–612. https://doi.org/10.1007/s00415-010-5799-6

Oliva, M., Berkovic, S. F., & Petrou, S. (2012). Sodium channels and the neurobiology of epilepsy: Sodium Channels and Epilepsy. Epilepsia, 53(11), 1849–1859. https://doi.org/10.1111/j.1528-1167.2012.03631.x

Paling, D., Golay, X., Wheeler-Kingshott, C., Kapoor, R., & Miller, D. (2011). Energy failure in multiple sclerosis and its investigation using MR techniques. Journal of Neurology, 258(12), 2113–2127. https://doi.org/10.1007/s00415-011-6117-7

Petroff, O. A. C., Errante, L. D., Kim, J. H., & Spencer, D. D. (2003). N-acetyl-aspartate, total creatine, and myo-inositol in the epileptogenic human hippocampus. Neurology, 60(10), 1646–1651. https://doi.org/10.1212/01.WNL.0000068020.85450.8B

Pike, M. M., Frazer, J. C., Dedrick, D. F., Ingwall, J. S., Allen, P. D., Springer, C. S., & Smith, T. W. (1985). 23Na and 39K nuclear magnetic resonance studies of perfused rat hearts. Discrimination of intra-and extracellular ions using a shift reagent. Biophysical Journal, 48(1), 159–173. https://doi.org/10.1016/S0006-3495(85)83769-3

Rajan, J., Poot, D., Junta, J., & Sijbers, J. (2010). Noise measurement from magnitude MRI using local estimates of variance and skewness. Physics in Medicine and Biology, 55(22), 6973–6973. https://doi.org/10.1088/0031-9155/55/22/6973

Reetz, K., Romanzetti, S., Dogan, I., Saß, C., Werner, C. J., Schiefer, J., Schulz, J. B., & Shah, N. J. (2012). Increased brain tissue sodium concentration in Huntington’s Disease—A sodium imaging study at 4T. NeuroImage, 63(1), 517–524. https://doi.org/10.1016/j.neuroimage.2012.07.009

Ridley, B., Marchi, A., Wirsich, J., Soulier, E., Confort-Gouny, S., Schad, L., Bartolomei, F., Ranjeva, J.-P., Guye, M., & Zaaraoui, W. (2017). Brain sodium MRI in human epilepsy: Disturbances of ionic homeostasis reflect the organization of pathological regions. NeuroImage, 157, 173–183. https://doi.org/10.1016/j.neuroimage.2017.06.011

Ridley, B., Nagel, A. M., Bydder, M., Maarouf, A., Stellmann, J.-P., Gherib, S., Verneuil, J., Viout, P., Guye, M., Ranjeva, J.-P., & Zaaraoui, W. (2018). Distribution of brain sodium long and short relaxation times and concentrations: A multi-echo ultra-high field 23Na MRI study. Scientific Reports, 8(1), 4357. https://doi.org/10.1038/s41598-018-22711-0

Rooney, W. D., & Springer, C. S. (1991). A comprehensive approach to the analysis and interpretation of the resonances of spins 3/2 from living systems. NMR in Biomedicine, 4(5), 209–226. https://doi.org/10.1002/nbm.1940040502

Rose, C. R., & Karus, C. (2013). Two sides of the same coin: Sodium homeostasis and signaling in astrocytes under physiological and pathophysiological conditions: Sodium Homeostasis in Astrocytes. Glia, 61(8), 1191–1205. https://doi.org/10.1002/glia.22492

Seabold, S., & Perktold, J. (2010). Statsmodels: Econometric and Statistical Modeling with Python. 92–96. https://doi.org/10.25080/Majora-92bf1922-011

Seifert, G., & Steinhäuser, C. (2013). Neuron–astrocyte signaling and epilepsy. Experimental Neurology, 244, 4–10. https://doi.org/10.1016/j.expneurol.2011.08.024

Simister, R. J., McLean, M. A., Barker, G. J., & Duncan, J. S. (2009). Proton MR spectroscopy of metabolite concentrations in temporal lobe epilepsy and effect of temporal lobe resection. Epilepsy Research, 83(2), 168–176. https://doi.org/10.1016/j.eplepsyres.2008.11.006

Simister, R. J., Woermann, F. G., McLean, M. A., Bartlett, P. A., Barker, G. J., & Duncan, J. S. (2002). A Short-echo-time Proton Magnetic Resonance Spectroscopic Imaging Study of Temporal Lobe Epilepsy. Epilepsia, 43(9), 1021–1031. https://doi.org/10.1046/j.1528-1157.2002.50701.x

Sofroniew, M. V., & Vinters, H. V. (2010). Astrocytes: Biology and pathology. Acta Neuropathologica, 119(1), 7–35. https://doi.org/10.1007/s00401-009-0619-8

Statistical Parametric Mapping: The Analysis of Functional Brain Images—1st Edition. (n.d.). Retrieved November 3, 2021, from https://www.elsevier.com/books/statistical-parametric-mapping-the-analysis-of-functional-brain-images/penny/978-0-12-372560-8

Stefano, N. D., Matthews, P. M., Ford, B., Genge, A., Karpati, G., & Arnold, D. L. (1995). Short-term dichloroacetate treatment improves indices of cerebral metabolism in patients with mitochondrial disorders. Neurology, 45(6), 1193–1198. https://doi.org/10.1212/WNL.45.6.1193

Stys, P., Waxman, S., & Ransom, B. (1992). Ionic mechanisms of anoxic injury in mammalian CNS white matter: Role of Na+ channels and Na(+)-Ca2+ exchanger. The Journal of Neuroscience, 12(2), 430–439. https://doi.org/10.1523/JNEUROSCI.12-02-00430.1992

Urenjak, J., Williams, S. R., Gadian, D. G., & Noble, M. (1993). Proton nuclear magnetic resonance spectroscopy unambiguously identifies different neural cell types. The Journal of Neuroscience: The Official Journal of the Society for Neuroscience, 13(3), 981–989.

Vagnozzi, R., Tavazzi, B., Signoretti, S., Amorini, A. M., Belli, A., Cimatti, M., Delfini, R., Di Pietro, V., Finocchiaro, A., & Lazzarino, G. (2007). TEMPORAL WINDOW OF METABOLIC BRAIN VULNERABILITY TO CONCUSSIONS. Neurosurgery, 61(2), 379–389. https://doi.org/10.1227/01.NEU.0000280002.41696.D8

van der Hel, W. S., van Eijsden, P., Bos, I. W. M., de Graaf, R. A., Behar, K. L., van Nieuwenhuizen, O., de Graan, P. N. E., & Braun, K. P. J. (2013). In vivo MRS and histochemistry of status epilepticus-induced hippocampal pathology in a juvenile model of temporal lobe epilepsy: STATUS EPILEPTICUS-INDUCED HIPPOCAMPAL PATHOLOGY. NMR in Biomedicine, 26(2), 132–140. https://doi.org/10.1002/nbm.2828

Voets, N. L., Hodgetts, C. J., Sen, A., Adcock, J. E., & Emir, U. (2017). Hippocampal MRS and subfield volumetry at 7T detects dysfunction not specific to seizure focus. Scientific Reports, 7(1), 16138. https://doi.org/10.1038/s41598-017-16046-5

Wang, Y., Majors, A., Najm, I., Xue, M., Comair, Y., Modic, M., & Ng, T. C. (1996). Postictal alteration of sodium content and apparent diffusion coefficient in epileptic rat brain induced by kainic acid. Epilepsia, 37(10), 1000–1006. https://doi.org/10.1111/j.1528-1157.1996.tb00539.x

Wang, Y.-H., An, Y., Fan, X.-T., Lu, J., Ren, L.-K., Wei, P.-H., Cui, B.-X., Du, J.-L., Lu, C., Wang, D., Zhang, H.-Q., Shan, Y.-Z., & Zhao, G.-G. (2018). Comparison between simultaneously acquired arterial spin labeling and 18F-FDG PET in mesial temporal lobe epilepsy assisted by a PET/MR system and SEEG. NeuroImage. Clinical, 19, 824–830. https://doi.org/10.1016/j.nicl.2018.06.008

Waxman, S. G. (2006). Axonal conduction and injury in multiple sclerosis: The role of sodium channels. Nature Reviews. Neuroscience, 7(12), 932–941. https://doi.org/10.1038/nrn2023

Zaaraoui, W., Konstandin, S., Audoin, B., Nagel, A. M., Rico, A., Malikova, I., Soulier, E., Viout, P., Confort-Gouny, S., Cozzone, P. J., Pelletier, J., Schad, L. R., & Ranjeva, J.-P. (2012). Distribution of brain sodium accumulation correlates with disability in multiple sclerosis: A cross-sectional 23Na MR imaging study. Radiology, 264(3), 859–867. https://doi.org/10.1148/radiol.12112680

Zhao, H., Carney, K. E., Falgoust, L., Pan, J. W., Sun, D., & Zhang, Z. (2016). Emerging roles of Na+/H+ exchangers in epilepsy and developmental brain disorders. Progress in Neurobiology, 138–140, 19–35. https://doi.org/10.1016/j.pneurobio.2016.02.002

