## Supplemental Figure 1 : Panel of spectra as explored during quality assessment. for "Combining Sodium MRI, Proton MR Spectroscopic Imaging and Intracerebral EEG in Epilepsy"

### Supplementary Material

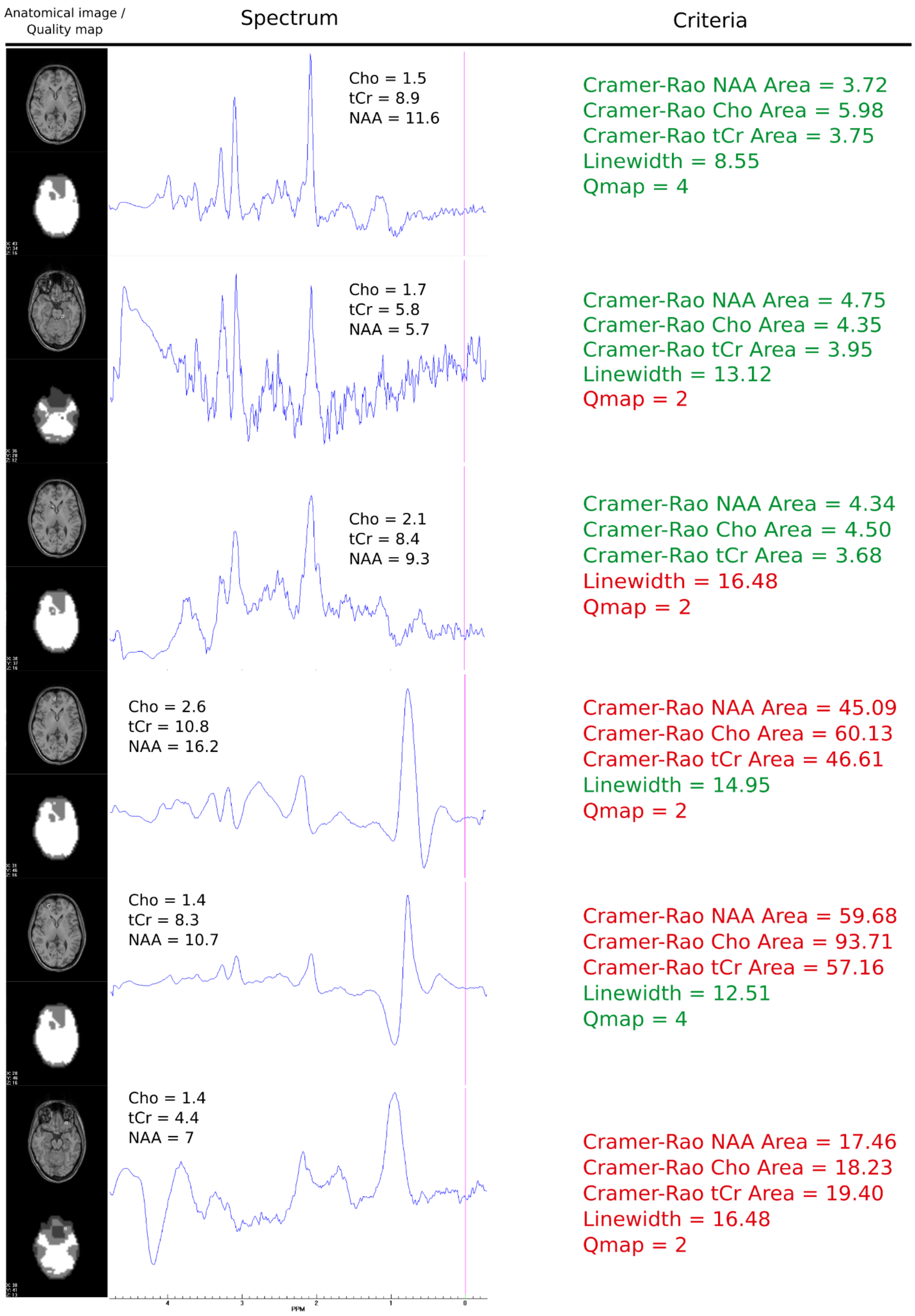


**Supplemental Figure 1 : Panel of spectra as explored during quality assessment.** Here are presented spectra and the associated Cramer-Rao Bound, linewidth and Qmap values in a healthy control subject. According to the selected thresholds, these values are in red when rejected and green otherwise. The absolute values of NAA, Cho and tCr are above the corresponding spectrum. On the left are represented the anatomical image with the selected voxel, providing the corresponding spectrum on the right, and the associated Qmap just underneath. We can observe spectrum deterioration as more and more criteria are in red.
