## Supplemental Table 1. Total number of ROIs per zone for "Combining Sodium MRI, Proton MR Spectroscopic Imaging and Intracerebral EEG in Epilepsy"

**Supplemental Table 1. Total number of ROIs per zone, with $^{23}$Na-MRI data ($n_{Patients}$ = 15, $n_{HC}$ = 18), with $^{1}$H-MRSI data ($n_{Patients}$ = 17, $n_{HC}$ = 25), and with both modalities ($n_{Patients}$ = 13)**

| | | **EZ** | **PZ** | **NIZ** | **Total** | **Mean** ± SD |
|---|---|---|---|---|---|---|
| **Na-MRI** | Patients ROIs | 107 | 149 | 483 | 739 | 49.27 ± 20.98 |
| | HC All ROIs | 1926 | 2682 | 8694 | 13302 | 886.8 ± 351.93 |
| **H-MRSI** | Patients ROIs | 77 | 134 | 477 | 688 | 40.47 ± 15.55 |
| | HC All ROIs | 1025 | 3350 | 11925 | 17200 | 1011.76 ± 388.82 |
| **Both** | Patients ROIs | 38 | 84 | 287 | 409 | 31.46 ± 16.64 |
