## Supplemental Table 2. ROIs 23Na-MRI and 1H-MRSI measures for "Combining Sodium MRI, Proton MR Spectroscopic Imaging and Intracerebral EEG in Epilepsy"

**Supplemental Table 2. ROIs $^{23}$Na-MRI and $^{1}$H-MRSI measures raw values mean, standard deviation and increase percentage compared to controls.**

| ROI | f | TSC (mM) | Cho (nAU) | NAA (nAU) | tCr (nAU) |
|---|---|---|---|---|---|
| **EZ** | 0.48 ± 0.18 (9.09%) | 58.12 ± 8.68 (3.73%) | 1.80 ± 0.55 (5.65%) | 7.95 ± 2.22 (-10.78%) | 6.87 ± 1.43 (-4.67%) |
| **PZ** | 0.5 ± 0.18 (4.17%) | 58.0 ± 9.08 (5.65%) | 1.65 ± 0.50 (2.51%) | 8.63 ± 2.46 (-6.84%) | 6.87 ± 1.54 (-3.32%) |
| **NI** | 0.51 ± 0.16 (2.00%) | 58.12 ± 10.56 (7.79%) | 1.54 ± 0.41 (-4.56%) | 9.00 ± 2.70 (-7.17%) | 6.82 ± 1.62 (-6.30%) |
| **All** | 0.51 ± 0.17 (4.08%) | 58.1 ± 10.01 (6.76%) | 1.59 ± 0.41 (-1.98%) | 8.81 ± 2.63 (-7.47%) | 6.83 ± 1.58 (-5.54%) |
